# Reemergence of Acute ischemic Stroke hospitalizations in the United States from 2010 to 2020

**DOI:** 10.1101/2023.06.25.23291876

**Authors:** Ahmed M. Elbayomy, Jason J Kim, Simon G Ammanuel, Edward L. Bradbury, Robert J Dempsey, Azam S. Ahmed

## Abstract

**Background and Objectives:** Stroke hospitalization rates in the United States are unequally distributed across the population. Factors such as age, race, socioeconomic status, and sex are associated with different hospitalization rates for acute ischemic stroke (AIS). This study aimed to review the patterns in these rates from 2010 to 2020.

**Methods:** The National Inpatient Sample (NIS) was used to analyze patterns in AIS hospitalizations in the United States between 2010 and 2020. The hospitalization rates were examined based on age, sex, race, region, stroke comorbidities, and income.

**Results:** The overall rate of AIS hospitalization increased from 230 to 254 per 100,000 individuals (+10.4%) between 2010 and 2020. Initially, the rates declined from 2010 to 2015 (230 to 227 per 100,000); however, from 2016 to 2020, the AIS hospitalization rates (AIS-HR) increased (242 to 254 per 100,000). The rate of AIS hospitalization increased significantly for individuals aged 25–44 years (25–37 per 100,000, +48.0%) and 45–65 years (171–235 per 100,000, +37.4%). AIS hospitalization rates remained relatively stable for individuals aged 65– 84 years (669–688 per 100,000, +3.7%) and declined for those over 85 years (2005–1756 per 100,000, -12.4%). Among sex comparisons, AIS hospitalization rates were stable in women (241–250 per 100,000, +3.7%) and increased in men (270–308 per 100,000, +14.1%). In terms of race, Blacks had the highest age-adjusted yearly hospitalization rate (437), followed by Whites (215) and Hispanics (207) per 100,000 in 2020. There was a stepwise increase in AIS hospitalization rates based on the Charlson Comorbidity Index (CCI), with White, Black, and Hispanic populations having AIS hospitalization rates of CCI 0 and 1 (533, 434, and 233), CCI 2 (2197, 2555, and 2125), and CCI ≥ 3 (5283, 5724, and 4767) per 100,000, respectively.

**Discussion:** Although the overall AIS hospitalization rate increased between 2010 and 2020, there was an initial decline until 2015, followed by a resurgence. This resurgence in AIS hospitalization rates was driven by greater underlying comorbidities, with disproportionate increases observed in Black and Hispanic populations. Further population analyses and cohort studies are required to confirm our findings.

## Introduction

Acute ischemic stroke (AIS) is one of the leading causes of death and disability in the United States, with a significant proportion of preventable cases.^1,2^ The likelihood of stroke can be decreased by controlling modifiable risk factors such as blood pressure and cholesterol, smoking cessation, and reducing sodium intake.^2^ However, these risk factors and the resulting rates of AIS are unevenly distributed among different racial/ethnic, socioeconomic, and age groups.^2-4^ Despite the recent emphasis on guideline-based care and the expectation that stroke hospitalization rates (HR) would decrease,^1,5^ the benefits of public health campaigns and improved access to primary care have not been equally experienced by all population segments. This is particularly evident among Black Americans, who have a 2-to 5-fold higher stroke rate than Whites, depending on age.^6^ An analysis of data from the National Inpatient Sample (NIS), the largest all-payer database in the US, consistently shows disparities in AIS-HR based on age, sex, and race.^3,4^

Although older adults account for most strokes, their AIS-HR patterns vary over time. Between 1988 and 2004, the AIS-HR steadily declined, with the most significant reduction observed in patients aged ≥ 65 years.^5^ Furthermore, between 1997 and 2006, the HR decreased in men and women aged 36–64.^1^ However, since then, hospitalization due to stroke has increased in younger patients, particularly in those aged 25–44.^7^ This finding is supported by a study reported a reduction in HR in patients aged > 65 years and an increased rate in individuals aged 25–64.^4^

Sex differences in AIS-HR have been identified, with men having a higher incidence than women. As individuals age, this pattern reverses, with a higher incidence.^8^ From 2000 to 2010, hospitalizations among women were lower and reduced more significantly than those among men.^4^ Between 2000 and 2010, age-adjusted hospitalizations increased for Blacks but decreased for Hispanics and Whites.^4^ Interestingly, one study demonstrated that Hispanics may have lower rates than Whites.^3^

Overall, numerous factors influence the distribution of AIS in the US population. These factors are dynamic and crucial for understanding differences in stroke rates among different population groups. Developing public health programs that address these disparities requires a comprehensive understanding of these factors. Therefore, our objective was to provide updated patterns in AIS-HR in the United States using the latest data (from 2010 to 2020) by age, race, and sex. We evaluated the underlying comorbidities and household income of affected individuals.

## Methods

### Study design

This retrospective longitudinal study examined patients with AIS admitted to acute care hospitals in the United States. The data for this study were extracted from the Healthcare Cost and Utilization Project’s National (Nationwide) Inpatient Sample (NIS) database, covering 2010 to 2020.^9^ The NIS database, maintained by the Agency for Healthcare Research and Quality, is the largest publicly available all-payer inpatient database in the United States. It includes data from approximately 7 to 8 million hospital stays annually, obtained from a stratified sample of 20% of non-federal US hospitals in participating states.

To ensure national representativeness, each discharge in the NIS was weighted using the following formula: weight = (total number of discharges from all acute care hospitals in the United States) / (number of discharges included in the 20% sample).^9^ This weighting approach allows researchers to generalize the findings to the entire population of acute care hospital discharges in the United States. The NIS supplies discharge weights (DISCWT) for the sampled discharges, which when applied to the sample provides a weighted national estimate. Prior to redesigning in 2012, the NIS was constructed by retaining 100% of the discharges from 20% of the randomly sampled community hospitals. In 2012, the NIS changed sampling strategy to include 20% discharges from all participating hospitals, to provide more precise estimates by reducing sampling error.

The NIS is a comprehensive source of hospital data that enables researchers to investigate various aspects of healthcare delivery and patient outcomes. It is a discharge-level database that contains de-identified clinical and non-clinical data elements at both patient and hospital levels. Consequently, multiple admissions for the same patient were treated as separate discharges and recorded individually in a database.^9^

Patient-level data encompassed various variables, including age, sex, race (categorized as White, Black, Hispanic, Asian or Pacific Islander, Native American, and others), median household income based on the patient’s zip code (divided into the 1^st^, 2^nd^, 3^rd^, and 4^th^ quartiles), primary expected payer (Medicare, Medicaid, private insurance, self-pay, no charge, and others), location (urban and rural), principal diagnosis, and secondary diagnosis. The diagnoses were coded using the International Classification of Diseases (ICD), 9^th^, and 10^th^ revisions.

To assess the comorbidity burden, Deyo’s modification of the Charlson comorbidity index (CCI) which is a method of categorizing comorbidities of patients based on ICD codes found in administrative data was used.^10^

Regarding ethical considerations, the study was exempted from review by the institutional review board because informed consent was not required as the participants’ identities were de-identified within the database.

The population estimates used in this study were obtained from the website of the United States Census Bureau.

### Selection criteria

Patients included in the analysis were ≥ 25 years of age and had a principal and secondary diagnosis of AIS. The specific diagnostic codes used were ICD-9-CM codes 433.x1, 434.x1, 436, and ICD-10-CM code I63.x and I64.^11^ For comorbidities and AIS related complications, we picked codes that have previously reported high sensitivity and specificity for these diagnoses.

Two denominators were used to calculate AIS admission rates: total hospital admissions and the general population in the United States. The rates of AIS hospitalization, broken down by age, sex, and race/ethnic group, were determined using the weighted number of hospitalizations as the numerator and the US civilian population as the denominator.

To facilitate comparisons between years and sexes, the annual rates were adjusted using the age distribution of the US female population in 2000. Similarly, for comparisons across years and races, rates were standardized using the age distribution of the US White population in 2000.

Because the data used in this study were population-based, the average rate of change was calculated as the average percentage change from the previous year across all years after 2010.^4^

We also looked at AIS-HR by comorbidities and COVID-19, COVID-19 was determined using code U07.1, which was only assigned to cases with confirmed laboratory evidence of SARS-COV-2 infection. As the U07.1 code became available in late March 2020, analyses utilizing this code were limited to the period of April to December 2020.

### Data analysis

All statistical analyses were conducted using STATA (version 17). The NIS database is based on a complex sampling design involving stratification, clustering, and weighting. STATA software ensured nationally representative and unbiased results and accurate variance estimates.

Patient-level observations were weighted to obtain estimates representing the entire population of hospitalized patients with AIS in the United States.

Fisher’s exact test was used to compare proportions, whereas the t-test was used to compare continuous variables between the two groups. Analysis of variance (ANOVA) was used to compare continuous variables between more than two groups.

The Cochran-Armitage Chi-square test for trends was used to analyze the AIS hospitalization rates for the years included in the study. In addition, the difference in incidence rate was calculated for each pair of consecutive study years to assess the rate of change in the incidence rate of AIS among the patient groups. Fisher’s exact test was used to compare the calculated differences in incidence rates between the two patient groups for each pair of consecutive study years.

## Results

### Study population

Table 1 (supplementary) summarizes patient characteristics. From 2010 to 2020, the number of AIS hospitalizations increased from 479,074 to 669,899 (Fig. 1). The mean age of the patients ranged from 69.5 to 70.9 years. Most patients were male (51.26%), identified as White (66.80%), and had insurance coverage through Medicare or private insurance (63.2% and 19.1%, respectively). Most patients had a comorbidity burden of CCI ≥ 3 (73.53%) and fell within the lowest third-income quartile (64.0%).

**Figure 1.**
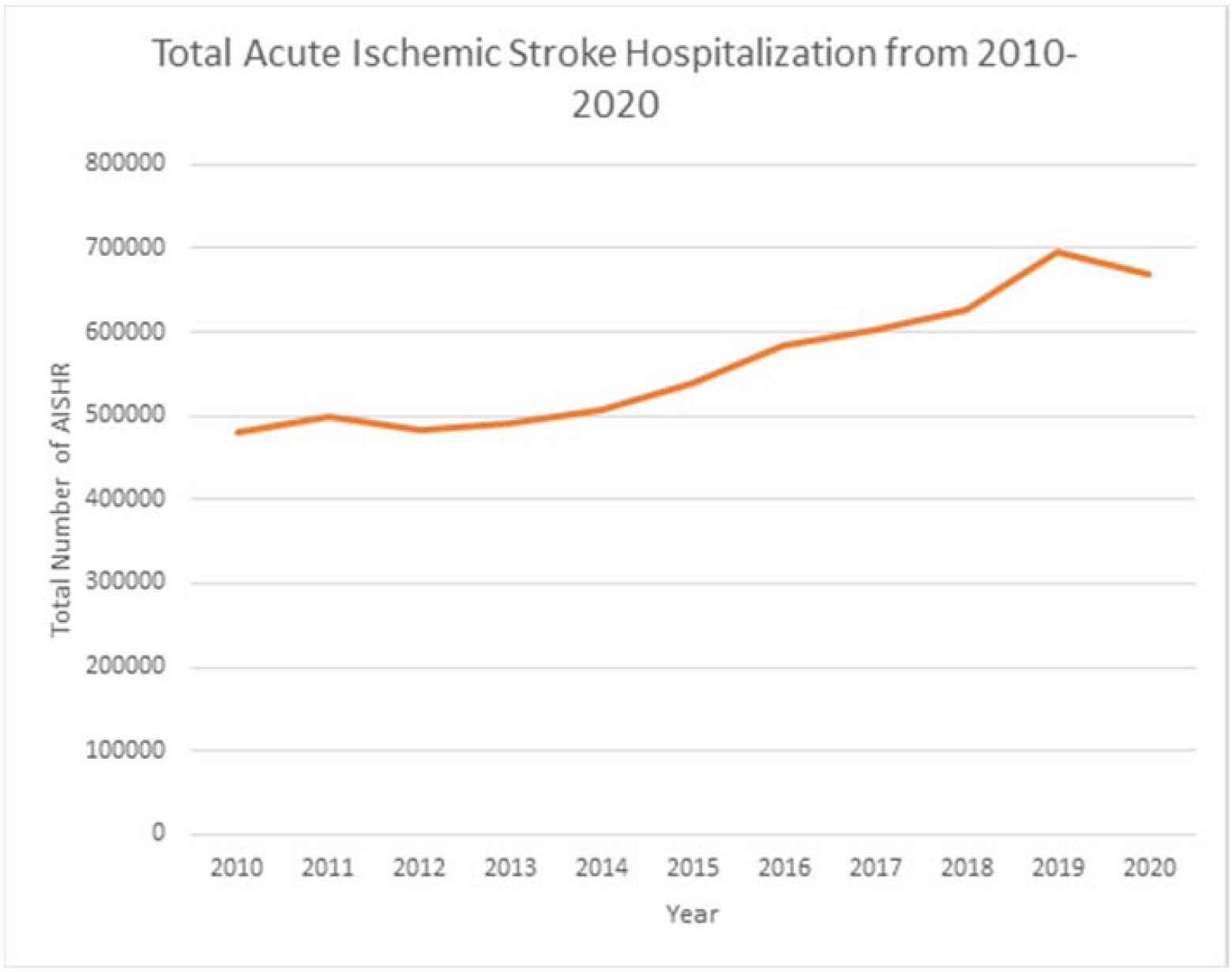

The AIS hospitalization rate in the United States increased from 230 to 254 per 100,000 people between 2010 and 2020, representing an overall rate increase of 10.4%. These rates were adjusted for age using 2000 US census data.

### General Patterns of AIS-HR

Table 2 provides an overview of the overall AIS-HR during the study period. The data revealed a modest decreasing trend in the AIS-HR from 2010 to 2015 (230–227 per 100,000 population). From 2016 to 2018, the rate remained relatively stable, ranging from 242 to 247 cases per 100,000 individuals. However, from 2018 onward, the AIS-HR sharply increased, reaching 254 per 100,000 by 2020 (p < 0.001) (Figure 1).

**Table 2:**

| 2010 | 2011 | 2012 | 2013 | 2014 | 2015 | 2016 | 2017 | 2018 | 2019 | 2020 | P for Trend |
| --- | --- | --- | --- | --- | --- | --- | --- | --- | --- | --- | --- |
| **Age-adjusted AIS hospitalization rates per 100.000 in the US population from 2010 to 2020** |  |  |  |  |  |  |  |  |  |  |  |
| 230 | 235 | 219 | 216 | 220 | 227 | 242 | 242 | 247 | 271 | 254 | <0.001 |
| **AIS hospitalization rates by age category per 100.000 in the US from 2010 to 2020** |  |  |  |  |  |  |  |  |  |  |  |
| **25 to 44** |  |  |  |  |  |  |  |  |  |  |  |
| 25<br>(24-26) | 24<br>(24-25) | 24<br>(23-25) | 25<br>(25-26) | 25<br>(25-26) | 28<br>(28-29) | 31<br>(31-32) | 32<br>(31-33) | 33<br>(33-34) | 37<br>(37-38) | 37<br>(36-38) | <0.001 |
| **45 to 64** |  |  |  |  |  |  |  |  |  |  |  |
| 171<br>(171-173) | 172<br>(171-173) | 169<br>(168-170) | 171<br>(170-172) | 177<br>(176-179) | 188<br>(187-190) | 201<br>(200-203) | 209<br>(208-210) | 216<br>(215-218) | 246<br>(245-247) | 235<br>(234-236) | <0.001 |
| **65 to 84** |  |  |  |  |  |  |  |  |  |  |  |
| 669<br>(667-673) | 696<br>(693-699) | 628<br>(626-631) | 616<br>(613-619) | 619<br>(617-622) | 636<br>(633-639) | 671<br>(668-673) | 669<br>(667-672) | 687<br>(684-690) | 730<br>(727-733) | 688<br>(686-691) | <0.001 |
| **85 and older** |  |  |  |  |  |  |  |  |  |  |  |
| 2005<br>(1992-2018) | 2024<br>(2011-2036) | 1890<br>(1878-1902) | 1799<br>(1788-1811) | 1842<br>(1831-1854) | 1804<br>(1792-1815) | 1947<br>(1935-1959) | 1860<br>(1849-1871) | 1786<br>(1775-1797) | 1999<br>(1987-2011) | 1756<br>(1746-1768) | <0.001 |
| **Race-specific age-adjusted AIS hospitalization rates per 100.000 in the US from 2010 to 2020** |  |  |  |  |  |  |  |  |  |  |  |
| **White** |  |  |  |  |  |  |  |  |  |  |  |
| 181 | 190 | 206 | 204 | 210 | 215 | 234 | 234 | 239 | 264 | 215 | <0.001 |
| **Black** |  |  |  |  |  |  |  |  |  |  |  |
| 406 | 386 | 355 | 347 | 355 | 385 | 399 | 413 | 412 | 464 | 437 | <0.001 |
| **Hispanic** |  |  |  |  |  |  |  |  |  |  |  |
| 182 | 204 | 189 | 191 | 184 | 190 | 197 | 208 | 222 | 219 | 207 | <0.001 |
| **Sex-specific age-adjusted AIS hospitalization rates per 100.000 in the US from 2010 to 2020** |  |  |  |  |  |  |  |  |  |  |  |
| **Males** |  |  |  |  |  |  |  |  |  |  |  |
| 270 | 272 | 255 | 254 | 261 | 271 | 288 | 289 | 297 | 326 | 308 | <0.001 |
| **Females** |  |  |  |  |  |  |  |  |  |  |  |
| 241 | 249 | 230 | 224 | 226 | 231 | 247 | 246 | 246 | 271 | 250 | <0.001 |

Examining age-specific AIS-HR from 2010 to 2020 (Table 2, Figure 2), it was found that individuals aged 25–44 years experienced an increase in AIS-HR from 25 to 37 per 100,000 (+48.0%), while those aged 45–64 years demonstrated an increase from 171 to 235 per 100,000 (+37.4%). In contrast, AIS-HR remained stable for individuals aged 65–84 years (669–688 per 100,000, +3.7%) and decreased for those aged 85 years and above (2005 to 1756 per 100,000, - 12.4%) (p < 0.0001) (Figure 2).

**Figure 2.**
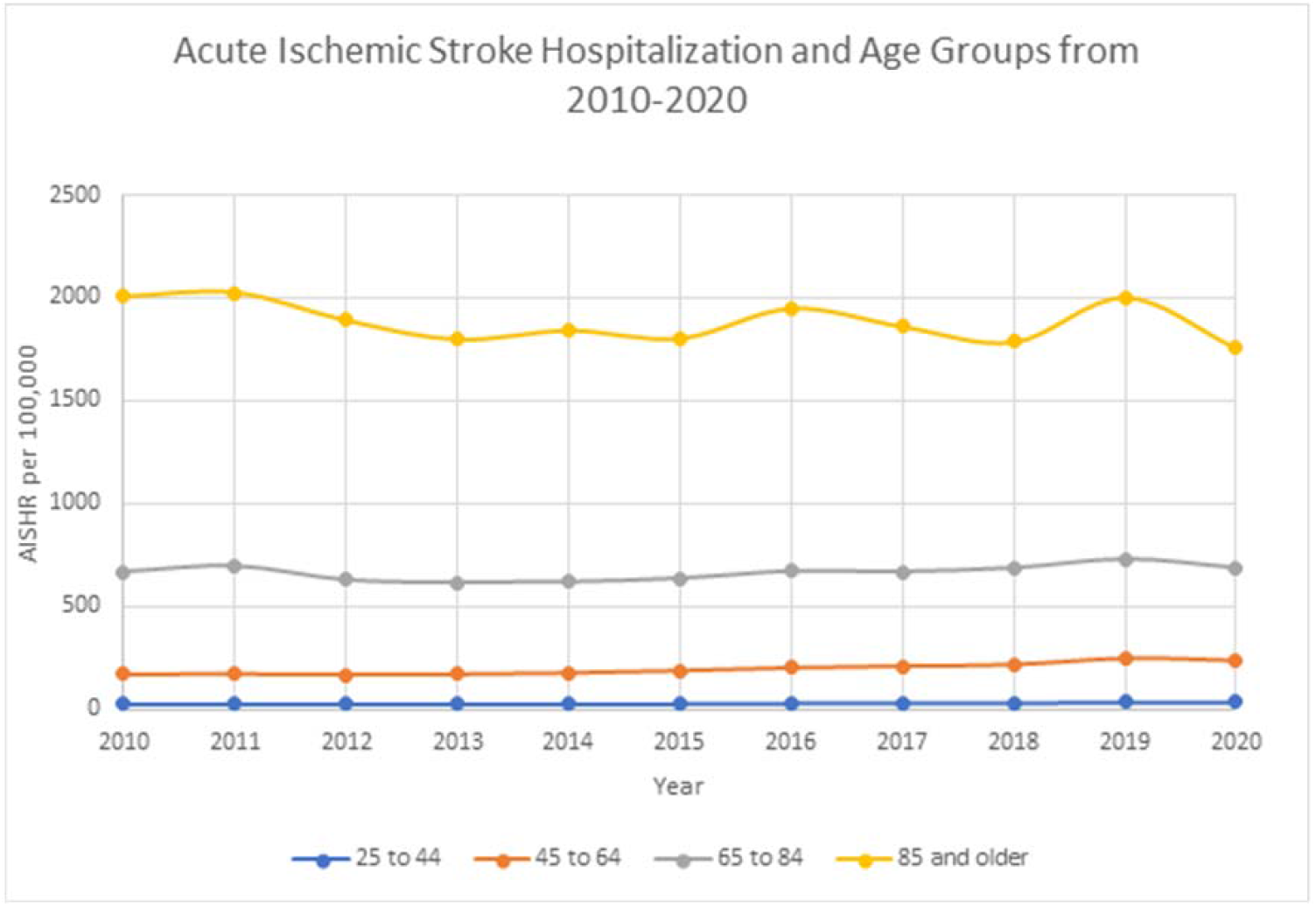

Furthermore, when analyzing age-adjusted AIS-HR by sex (Table 2, Figure 3), it was observed that men had higher rates than women. Men experienced a higher increase in AIS-HR (270–308 per 100,000, +14.1%) than did women (241–250 per 100,000, +3.7%) (p < 0.0001).

**Figure 3.**
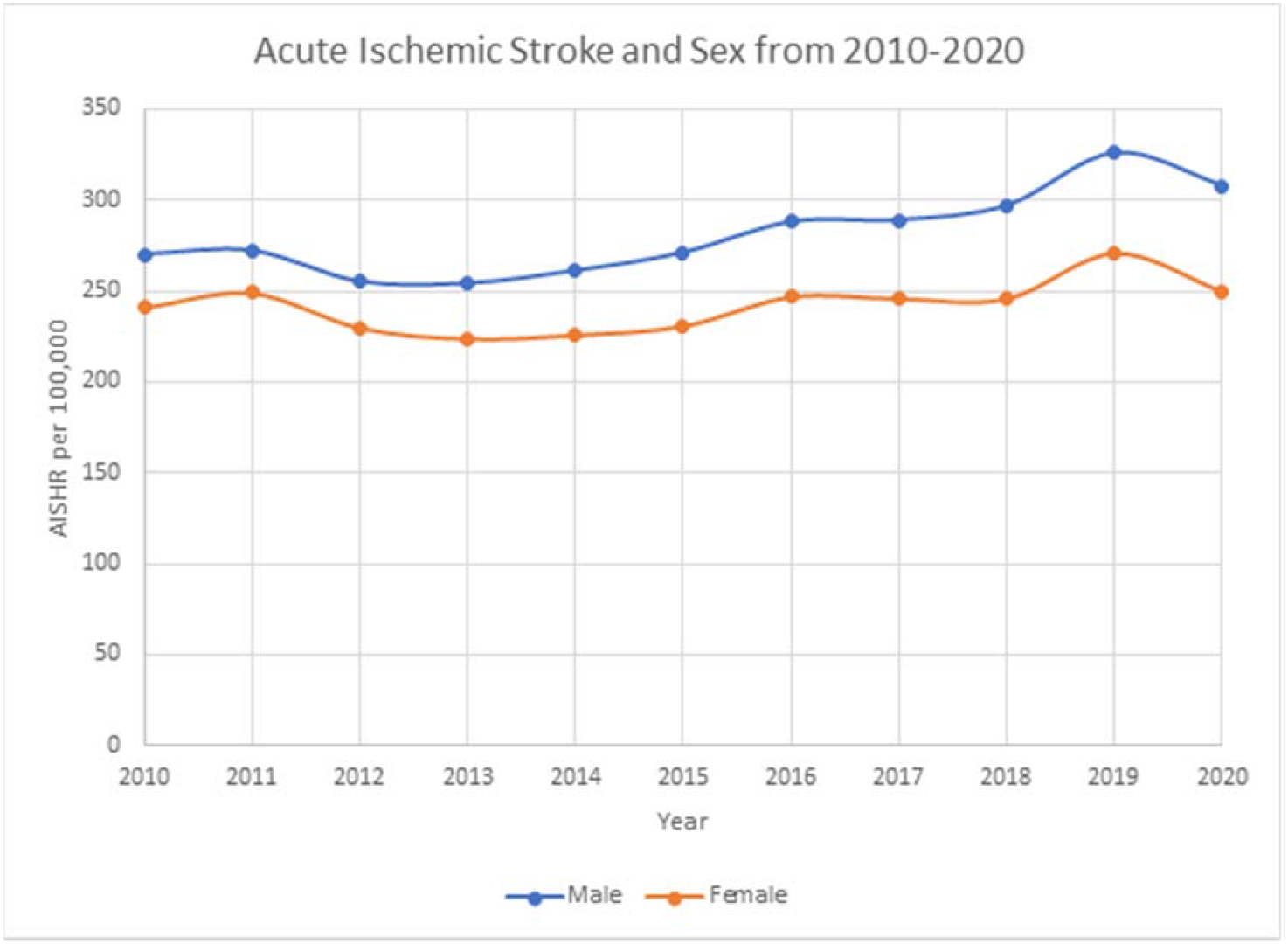

Regarding race/ethnicity, Blacks had the highest age-adjusted yearly hospitalization rates for AIS (437), followed by Whites (215), and Hispanics (207) in 2020. From 2010 to 2020, AIS-HR increased for Blacks (406–437), Whites (181–215), and Hispanics (182–207) (Figure 4).

**Figure 4.**
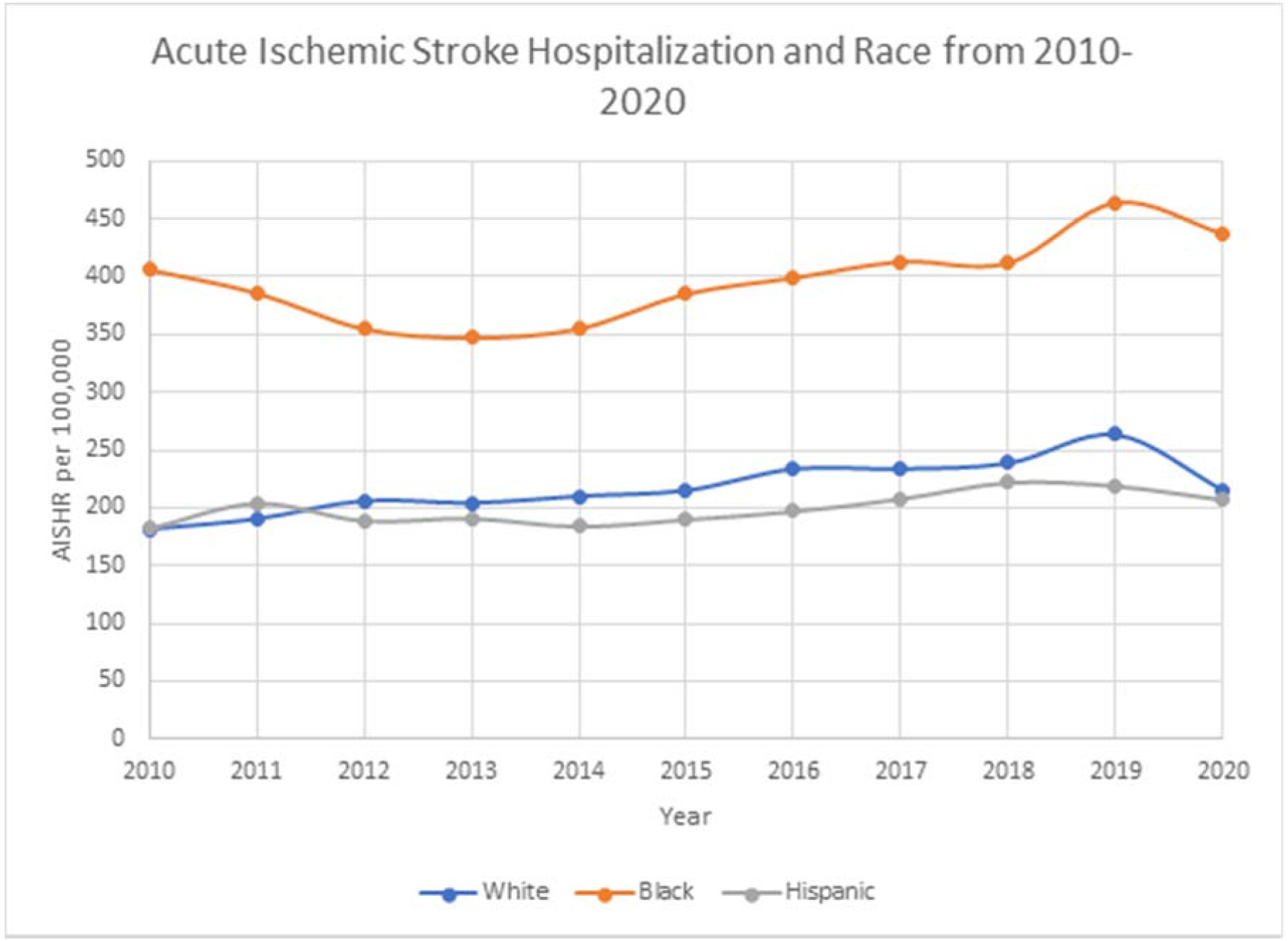

### Pattern of AIS-HR by Age, Race, and Comorbidities

As demonstrated by CCI, AIS-HR exhibited a general decline in cases with CCI < 3, whereas there was an increase in cases with CCI ≥ 3 between 2010 and 2020. When examining CCI by age, an increase was observed across all age groups, with the highest rate recorded in individuals aged ≥ 85 years, specifically in the CCI ≥ 3 categories (5379 vs. 6445 per 100,000) during that period (Table 3, Supplementary). Analysis of AIS-HR by race (Table 4, Supplementary) revealed that the CCI 3 or greater category exhibited the highest rate within each racial group, with Blacks experiencing the highest overall AIS-HR within the highest comorbidity categories (White: 3635 to 5283, Black: 3772–5724, Hispanic: 2862–4767 per 100,000). In summary, there was an overall increase in the AIS-HR across all comorbidity categories and races, with the greatest increase observed in the highest categories between 2010 and 2020.

### Patterns of AIS-HR by income, location, and hospital size

Analysis of AIS-HR by hospital size revealed a notable increase in the proportion of AIS-HR cases in small- and medium-sized hospitals, with percentages increasing from 11.37% to 18.93% and 21.43% to 27.77%, respectively, from 2010 to 2020 (p < 0.001). Conversely, rural hospitals experienced a decrease in AIS-HR cases compared to urban hospitals, with percentages declining from 12.74% to 7.03% and 87.26% to 92.97%, respectively (p < 0.001). Furthermore, teaching hospitals witnessed a progressive increase in AIS-HR cases from 48.13% to 76.46% between 2010 and 2020. By the end of 2020, teaching hospitals surpassed non-teaching hospitals regarding AIS-HR cases, with 51.87% and 23.54%, respectively (p < 0.001).

In terms of median household income, the highest quartile experienced a decrease in the proportion of AIS-HR cases, with percentages declining from 19.83% to 18.73%. The third quartile also witnessed a decrease from 23.55% to 22.63%. Conversely, the proportions increased in the first quartile (30.56%–31.14%) and the second quartile (26.06%–27.49%). However, these changes were not statistically significant (p = 0.14) (Table 1 supplementary).

### AIS-HR by comorbidities and COVID-19

A total of 674,354 patients with AIS and 1,678,995 patients with COVID-19 having primary or secondary diagnoses were identified in the NIS 2020 dataset. Among the 1,058,815 patients diagnosed with COVID-19 as their primary diagnosis, 7,560 (0.75%) developed AIS, whereas, among the 30,676,832 patients without COVID-19, 505,884 (1.64%) developed AIS. COVID-19 patients who developed AIS had a greater mean age than those who did not (68.74 vs. 64.32; p < 0.0001).

There was a lower proportion of White patients among patients with COVID-19 who developed AIS compared to those without stroke (44.91% vs. 50.09%; p < 0.0001), while the proportion of Black patients was higher (22.88% vs. 17.89%; p < 0.0001). However, there was no significant difference in the proportion of Hispanic patients between the two groups (18.19% vs. 20.06%; p = 0.06).

The proportions of patients with CCI ≥ 3 (68.32% vs. 27.54%; p < 0.0001), hypertension (74.47% vs. 67.11%; p < 0.0001), diabetes (40.94% vs. 34.18%; p < 0.0001), hyperlipidemia (41.79% vs. 39.02%; p = 0.02), atrial fibrillation (27.58% vs. 14.99%; p < 0.0001), coagulopathy (26.32% vs. 10.86%; p < 0.0001), and heart failure (23.54% vs. 15.62%; p < 0.0001) were higher among patients with COVID-19 who developed AIS than among those who did not.

## Discussion

This study used the NIS dataset to describe patterns in AIS rates from 2010 to 2020. Several patterns and characteristics observed since 2000 have persisted, along with new insights and changes in the growing population and overall health of the United States.

The results of this study indicated that age-adjusted AIS-HR remained relatively stable from 2010 to 2020, despite a noticeable increase in the rate over the past four years. This contrasts with a study, which covered the period from 2000 to 2010 and showed a decrease in the AIS-HR. Our study revealed a relatively constant rate until the resurgence of stroke rates after 2017.^4^ These findings are consistent with those of Ramphul et al., who suggested the possibility of a resurgence in AIS-HR beyond 2010.^3^

From 2010 to 2020, stroke incidence increased in all age groups. Among these age groups, individuals aged 65–84 years accounted for most AIS hospitalizations, followed by those aged 45–64 years and those over 85 years. We observed an overall increase in AIS hospitalizations among individuals aged 25–64 years after 2017, with an increase in all age categories. These findings are consistent with recent research and previous studies using NIS, which also reported increased stroke incidence in younger individuals.^4,6,7^

The association between obesity and stroke risk is particularly interesting because obesity rates have increased among adults.^12,13^ This increase has contributed to higher stroke risk, especially in younger age groups. Younger individuals experience higher rates of AIS comorbidities such as hypertension and hyperlipidemia.^14,15^ While stroke in younger patients was previously attributed to trauma, the growing prevalence of obesity, diabetes, and hypertension significantly increased AIS in younger age groups.^14^

Our study reflects these patterns. Initially, most AIS hospitalizations in the 25-44 age group were shared by individuals in the lowest and highest comorbidity categories. However, in the decade leading up to 2020, the highest comorbidity category gradually became the predominant contributor to AIS hospitalizations in this age group.

Regarding differences in AIS-HR between sexes, our findings are consistent with previous literature, showing that males have a higher overall incidence of AIS-HR than females.^3^ However, in the latter half of the decade, women and men experienced increased AIS hospitalizations, which may be attributed to a general increase in AIS-HR cases. A study demonstrated that AIS-HR in women may have increased throughout the decade.^3^ This could be due to the increased use of birth control, higher prevalence of migraines and pregnancies, and an increase in obesity, hypertension (HTN), and high lipid levels (HLD) in the general population. These factors may contribute to a higher risk in women than previously thought.^14,16^ However, this increase may also be explained by a lack of awareness, insufficient education, and policy changes regarding the disparity in diagnosis and treatment between sexes.^17^

Regarding the effect of race on AIS-HR, our study revealed that Blacks had the highest rate of AIS hospitalization, followed by Whites and Hispanics. This finding aligns with the results of a previous study.^4^ However, when adjusting for age, the overall age-adjusted AIS-HR in Whites increased from 2010 to 2020, while other racial groups experienced a significant increase after 2017. This contradicts Ramierez et al.’s findings from 2000 to 2010, which showed a consistent pattern of increased AIS-HR among Blacks and reduced rates among Whites and Hispanics.^4^ While our results align with the fact that Black individuals still have the highest AIS-HR, we observed a gradual increase in AIS hospitalizations among the white population, and a delayed, sharp increase in the Black and Hispanic populations.

Although the reasons for the observed delay are not clearly understood, they are likely influenced by multiple factors that work synergistically. These factors may include suspicion toward the healthcare system among minority populations and systemic racism, leading to a higher likelihood of delayed presentation of acute stroke symptoms and chronic conditions such as diabetes.^18-20^ Moreover, the overall disparity in AIS rates among Black individuals may be attributed to the higher prevalence of comorbidities and differences in socioeconomic status within their communities.^3,21^ However, it is essential to note that recent studies have only accounted for approximately 50% of the observed effects, indicating that more research is needed to fully understand the underlying causes.^21^

For Hispanics, AIS and mortality rates were underestimated. This can be attributed to the method used in the NIS database, where race and ethnicity are combined, and limitations in the census data collection.^4^ Therefore, studying the racial effect on AIS using standard demographic data is challenging, as racial populations are not homogeneous, and essential aspects of race, community, and ethnicity remain unaccounted for. Nonetheless, the intersection of high-risk demographic categories leads to a particularly unfavorable synergistic effect. For example, Black individuals with a CCI score of 3 or higher had a 20% higher AIS-HR than Whites in the same category from 2010 to 2020.

Most patients in our study received treatment at large or medium-sized academic centers. Additionally, we observed that the stroke rate was higher in patients residing in the 1^st^ and 3^rd^ quartile of median house income. In contrast, it was lower in zip codes within the 2^nd^ and 4^th^ quartile of median house income. These findings suggest that access to healthcare services may present a barrier for individuals residing in less populated areas or those with limited financial resources.

Interestingly, we also noticed an increase in AIS hospitalizations in small and medium-sized hospitals. Simultaneously, the number of large hospitals decreased from 2010 to 2020. However, there has been a pattern toward rural and non-teaching hospitals admitting fewer patients for AIS hospitalization. This observation aligns with the findings of a previous study, which demonstrated that rural hospitals were less likely to administer tissue plasminogen activators (tPA) or provide endovascular therapy based on data from the NIS between 2012 and 2017.^22^ This pattern appears to have persisted since 2004, as another study revealed an increasing disparity in tPA usage.^23^ Therefore, our observations suggest that, although more small and medium-sized hospitals treat stroke patients, they are predominantly located in populated areas. Meanwhile, there is a growing lack of access to stroke care in rural and low-income populations.

The reasons for the overall increase in the AIS-HR after 2017 remain unclear. However, we observed a decrease in the lowest comorbidity category among those who experienced AIS hospitalizations, whereas the highest comorbidity category (CCI ≥ 3) showed an increase. This pattern may be attributed to public health interventions and guideline-based approaches to preventing stroke and other vascular diseases. These efforts have decreased the overall stroke rate, except in the most severe cases.^24,25^ Comorbidities play a significant role in driving AIS-HR. For example, regardless of race, the AIS-HR increased 4- to 5-fold when the CCI score increased from 2 to 3 or higher. These findings and those of other studies have led to the development of resources and interventions that target those at the highest risk.

National programs such as WISEWOMAN (Well-Integrated Screening and Evaluation for Women Across the Nation), the Paul Coverdell National Acute Stroke Program, and Million Hearts have been implemented to address disparities and focus on reducing risk factors. Some programs specifically emphasize the health of high-risk demographics. State and local initiatives have also been implemented, such as the Wisconsin Chronic Disease Prevention Program and the Oneida Stroke Prevention Program in Wisconsin.

Unsurprisingly, we have not yet witnessed a significant effect of these programs, considering the potentially long timeframe required for risk factor modification to manifest as reduced stroke rates. Additionally, access to these programs and preventative care is often limited among populations in which AIS-HR is most prevalent, such as low-income and minority communities. Distrust in healthcare systems and low health literacy among patients further contribute to this negative synergy. Therefore, it is crucial to maintain a sustained focus on prevention and stroke risk factor modification, with particular attention given to individuals with higher comorbidity burdens and those residing in low-income and minority communities.

From 2019 to 2020, the world experienced the COVID-19 pandemic, which introduced specific patterns and potential effects that may be reflected in our study. Many elective procedures were canceled, and treatment protocols and healthcare capacity were altered. Although we adjusted for COVID-19 diagnosis and other comorbidities in our analysis, numerous unmeasurable variables could have significantly influenced AIS-HR. Our study observed significant differences in stroke comorbidities between patients with and without COVID-19. The increased prevalence of comorbidities among patients with COVID-19 may contribute to a higher likelihood of intervention and hospital admission, thereby increasing the AIS-HR. This hypothesis is consistent with previous studies that examined the association between COVID-19 and stroke.^26-28^ It has been shown that although stroke is relatively rare among patients with COVID-19, the risk increases when individuals have cardiovascular risk factors such as hypertension, diabetes, hyperlipidemia, atrial fibrillation, and congestive heart failure.^28^ It has been highlighted the interplay between various mechanisms in COVID-19, including the development of cytokine storms, activation of the innate immune system, hypoxia-induced ischemia, and endothelial injury, which can affect pre-existing comorbidities.^27^ Additionally, Tu et al. demonstrated that this increased risk might occur in individuals with asymptomatic COVID-19.^29^

## Conclusion

In our observational study, which spanned from 2010 to 2020, we noted a resurgence in AIS-HR during the latter half of the decade. This increase was particularly evident in younger populations and women, although racial disparities in stroke rates persisted. Furthermore, individuals with multiple medical risk factors faced a higher and increasing burden of stroke, whereas healthy individuals had lower rates. Additionally, we observed a growing disparity in access to stroke treatment in low-income and rural regions. Based on our observations, the resurgence of AIS-HR may be attributed to the compounding interplay of multiple risk factors and reduced access to stroke treatment in various populations.

## Limitations

Our study had several limitations. First, the NIS used in our analysis is an inpatient database, meaning that it only captures data on patients admitted to the hospital. Therefore, the reported stroke rates in our study may have underestimated the overall stroke rate in the US. In addition, our study was based on observational data; therefore, we could only establish associations and formulate hypotheses based on our analysis. However, the analysis covered multiple years, allowing us to identify patterns in the data.

It is important to note that the dataset relied on the coding of hospitalization events, which could vary due to differences in hospital policies, provider preferences, and coding errors. Consequently, it is possible that not all cases of AIS hospitalizations were accurately captured in the dataset because coding practices may vary across different hospitals and providers. Moreover, our study encountered a transition point after 2015 when the International Classification of Diseases, Ninth Revision (ICD-9) codes were replaced with more complex and subcategorized ICD-10 codes. As a result, there may be specific codes that our study failed to capture from 2016 onward. Additionally, coding errors might have occurred when classifying transient ischemic attacks as AIS for ICD-9 and, potentially, ICD-10 codes.^11^ However, despite these limitations, the overall patterns in our data should remain consistent because we used all known codes equally across all years following the implementation of ICD-10 codes.

Furthermore, the potential influence of the COVID-19 pandemic on our results should be considered. The pandemic may have affected hospital protocols and capacities for stroke treatment and healthcare-seeking behavior in the general population. Therefore, 2019–2020 may have confounding effects from COVID-19 and the societal repercussions resulting from the pandemic. Interestingly, despite possible changes in access and capacity, we observed a sustained increase in AIS hospitalizations from 2019 to 2020.

Despite these limitations, our study benefited from its large sample size and national representation, which enhanced its generalizability to the entire nation. Our findings demonstrate an overall increase in stroke rates since 2010, accompanied by persistent disparities among racial demographics. Future research should focus on closely monitoring health interventions, changing treatment guidelines, and developing strategies to address existing health inequalities in our society.

## Data Availability

All data underlying the results are available as part of the article.

